# Short-Term Androgen Deprivation Therapy and ^18^F-flotufolastat PSMA PET/CT Imaging for Prostate Cancer Staging: A Pilot Study

**DOI:** 10.1101/2025.10.08.25337428

**Authors:** Pat F. Fulgham, Patrick W. Barr, Rodney R. Bowman, Gregory R. Thoreson, Stephanie Berger, Alexia Demitsas, Angela R. Clark

## Abstract

**Introduction:** This pilot study evaluates the impact of short-term androgen deprivation therapy (ADT) on the findings on ^18^F-flotufolastat PSMA PET/CT scan when used for staging prostate cancer prior to a robot-assisted radical prostatectomy (RARP) and bilateral pelvic lymphadenectomy (PLND).

**Methods:** Ten patients with unfavorable intermediate, high, or very-high risk prostate cancer were assigned to one of two cohorts, A and B (n=5 each). Baseline serum prostate-specific antigen (PSA) and testosterone were measured, followed by an ^18^F-flotufolastat PSMA PET/CT. All patients then underwent either 3 weeks (Cohort A) or 6 weeks (Cohort B) of relugolix therapy. A second PSMA PET/CT was performed immediately upon ADT completion. Quantitative and qualitative changes in PSMA PET/CT findings were analyzed.

**Results:** All patients reached castrate levels of testosterone (<20 ng/dL) within 3 weeks of the initiation of ADT. Only 1 patient (10%) exhibited a 16.1% increase in the maximum standardized uptake value (SUVmax) of the primary intraprostatic lesion. No new clinically significant findings were detected on the post-ADT PSMA PET/CT compared to pre-ADT scans, and no pre-existing findings were obscured post-ADT. No serious adverse events related to ADT were reported.

**Discussion:** Short-term ADT (3 and 6 weeks) did not alter PSMA PET/CT findings in a manner that would impact surgical planning or management.

## Introduction

The advent of PSMA PET/CT imaging has improved the specificity and sensitivity of imaging studies for the preoperative staging of patients prior to robot-assisted radical prostatectomy (RARP) compared to conventional imaging [1]. While PSMA PET/CT offers high sensitivity (85-97%) for detecting primary prostate cancer lesions, the sensitivity for detecting nodal metastasis before RARP remains low (40%) [2]. Approximately 13% of patients undergoing RARP for attempted cure of apparently localized disease will have evidence of biochemical recurrence within 10 years [3], highlighting the need for improved information about the presence of advanced localized or metastatic disease. More accurate staging would better inform the initial treatment choices and potential need for adjuvant or neoadjuvant therapy.

ADT may enhance the sensitivity of PSMA-PET/CT by inducing a flare in PSMA expression on prostate cancer cells in some patients. In 2018, Aggarwal et al [4] reported on 4 patients with metastatic castrate sensitive prostate cancer who underwent ADT prior to PSMA-PET/CT. PSMA expression increased in 3 of 4 patients with an average increase in SUVmax (Standardized Uptake Value) of 46.3% in the lesions which showed a response. Most of the responses occurred within 1 to 5 weeks. Additional work by Emmett et al [5], showed that the SUVmax increased in 3 of 8 patients (37.5%) with newly diagnosed metastatic castrate sensitive prostate cancer at 9 to 28 days after the initiation of ADT. The median increase in SUVmax for the patients who demonstrated an increase was 45%. Based on these studies, we hypothesized that short-term ADT could increase PSMA expression, potentially enhancing PSMA-PET/CT staging prior to RARP. We chose 3- and 6-week ADT durations to capture potential peak PSMA expression, while minimizing side effects from ADT, and to accommodate the common timing between diagnosis and surgery.

This pilot study investigates the effect of short-term ADT on the qualitative and quantitative findings of PSMA PET/CT for patients undergoing RARP.

## Materials and Methods

This prospective study was approved by the Institutional Review Board (IRB) and was performed in compliance with the Health Insurance Portability and Accountability Act (HIPAA). Written informed consent was obtained from all participants. This study was retrospectively registered on http://www.clinicaltrials.gov (NCT07069465).

A total of 10 patients with unfavorable intermediate, high, or very-high risk prostate cancer, scheduled to undergo RARP and standard pelvic lymph node dissection (PLND), were assigned to Cohort A (5 patients) or Cohort B (5 patients). Patients were assigned to Cohorts A or B based on order of presentation and the timing of intended RARP. All patients underwent a baseline PSMA PET/CT scan prior to initiation of relugolix (gonadotropin-releasing hormone receptor antagonist). Cohort A patients were treated with 3 weeks of relugolix followed by repeat PSMA PET/CT scan upon completion of relugolix. Cohort B patients were treated with 6 weeks of relugolix followed by repeat PSMA PET/CT upon completion of relugolix. All patients underwent RARP with PLND within 20 days (average 8.6 days, range 3-20 days) of completing the study protocol. Preoperative biopsy pathology and clinical stage were compared to surgical pathology and stage. A baseline serum prostate-specific antigen (PSA) and testosterone were obtained before the initiation of ADT, then at 3 weeks after the initiation of ADT, and at 6 weeks after the initiation of ADT (Fig.1).

**FIGURE 1.**
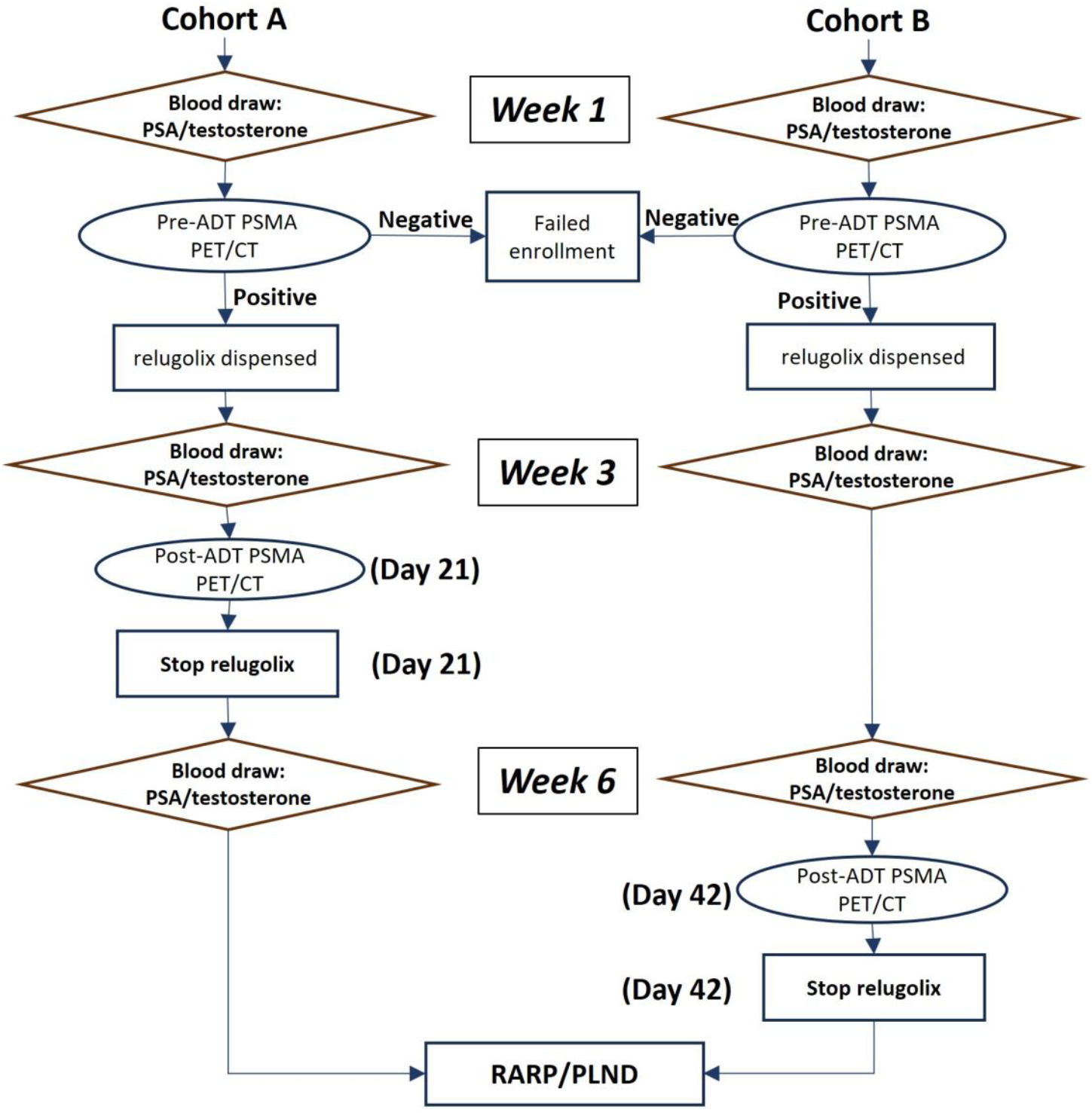
Study protocol for serum PSA, testosterone, and PSMA PET/CT testing following assignation to a cohort

PSMA PET/CT scans were obtained on a volumetric PET/CT system (GE Discovery VCT) with imaging performed at 60 minutes post infusion of ^18^F-flotufolastat PSMA PET/CT (Posluma®, Blue Earth Diagnostics). CT parameters included 3 mm slice thickness soft-tissue reconstruction kernel, 120 keV and 50 mAs, 600 mm field of view and a 512 matrix. ^18^F-flotufolastat is an (18)F radiopharmaceutical which is used as a radioactive diagnostic agent for positron emission tomography (PET) of PSMA positive lesions in men with prostate cancer [6]. Patients were injected with an average of 8.0 mCi (range 7.4 to 9.6 mCi) of ^18^F-flotufolastat. PET image reconstruction was performed using GE Discovery STE software.

PSMA PET/CT scans were evaluated by two nuclear medicine radiologists (Rad1 and Rad2). They did not have access to patient identification or sequence of the PSMA PET/CT scans and were blinded to the clinical outcomes. SUVmax was determined for the most prominent intra-prostatic lesion (henceforth referred to as the index lesion) as determined by the radiologist. Rad1 and Rad2 used MIM® Software version 7.3.6 (MIM Software, Inc., Cleveland, OH, USA). Liver background SUVmean and salivary uptake SUVmean were provided by Rad1 and Rad2 for each study. The radiologists assigned a PROMISE score (Prostate Cancer Molecular Imaging Standardized Evaluation) to each PSMA PET/CT scan as described by Eiber et al [7]. The miTNM score (molecular imaging, tumor, nodes, metastases) is a standardized reporting framework similar to the TNM classification for pathology.

## Results

The characteristics of individual patients as well as the average values and modes, where appropriate, for each cohort are summarized in Table 1. Age, testosterone, biopsy Gleason score, percentage of positive cores, grade group, and TNM (tumor, nodes, metastasis) clinical stage are comparable for Cohorts A and B.

**Table 1:** Patient Characteristics by Cohort.

| Cohort | Pt ID | Age Group | Baseline PSA (ng/mL) | Baseline Testosterone (ng/dL) | Biopsy Gleason Score | Biopsy Cores %Positive (# cores) | Grade Group | *cTNM |
| --- | --- | --- | --- | --- | --- | --- | --- | --- |
| A | 1 | 56-60 | 10.1 | 461 | 4+3 | 100% (12/12) | 3 | cT1cN0M0 |
| A | 2 | 66-70 | 8.1 | 574 | 4+4 | 42% (5/12) | 4 | cT1cN0M0 |
| A | 7 | 56-60 | 11.1 | 464 | 4+4 | 42% (5/12) | 4 | cT2bN0M0 |
| A | 9 | 56-60 | 5.6 | 439 | 4+3 | 43% (6/14) | 3 | cT1cN0M0 |
| A | 10 | 61-65 | 120.0 | 486 | 4+4 | 92% (11/12) | 4 | cT2cN0M0 |
| **A Avg (Range)** |  | **60 (56-67)** | **31.0 (5.6-120)** | **485 (439-574)** | **4+4 (mode)** | **63% (5-12)** | **4 (mode)** | **cT1cN0M0 (mode)** |
| B | 3 | 56-60 | 7.2 | 512 | 4+3 | 92% (11/12) | 3 | cT1cN0M0 |
| B | 4 | 66-70 | 11.8 | 485 | 4+4 | 17% (2/12) | 4 | cT1cN0M0 |
| B | 5 | 61-65 | 8.3 | 389 | 4+4 | 25% (3/12) | 4 | cT1cN0M0 |
| B | 6 | 66-70 | 7.3 | 453 | 4+4 | 92% (11/12) | 4 | cT3bN1M0 |
| B | 8 | 56-60 | 39.4 | 160 | 4+4 | 33% (4/12) | 4 | cT1cN0M0 |
| **B Avg (Range)** |  | **61.6 (57-66)** | **14.8 (7.2-39.4)** | **400 (160-512)** | **4+4 (mode)** | **52% (2-11)** | **4 (mode)** | **cT1cN0M0 (mode)** |

\*cTNM = Clinical tumor, node, metastasis stage

The disparity in baseline PSA average between Cohorts A and B is due to two outliers (one patient in Cohort A had PSA of 120.0 ng/mL and one patient in Cohort B had a PSA of 39.4 ng/mL). Excluding these two values the average baseline PSA was 8.7 ng/mL for both Cohort A and Cohort B. A serum testosterone of >150 ng/dL was documented for all patients prior to the initiation of ADT. The sample size in this pilot study is too small to permit reliable statistical comparisons.

The laboratory data related to ADT is summarized in Table 2. Nine of ten (90%) patients demonstrated a decline in PSA after 3 weeks of ADT (avg −69.6%). All patients reached castrate levels of testosterone (< 20 ng/dL).

**Table 2:** Laboratory data for individual patients and by Cohort, before and after ADT.

| Cohort | Pt ID | Baseline PSA (ng/mL) | Baseline Testosterone (ng/dL) | relugolix Duration | 2nd PSA (ng/mL) 3 weeks | 2nd Testosterone (ng/dL) 3 weeks | 3rd PSA (ng/mL) 6 weeks | 3rd Testosterone (ng/dL) 6 weeks |
| --- | --- | --- | --- | --- | --- | --- | --- | --- |
| A | 1 | 10.1 | 461 | 3 weeks | 4.6 | 9 | 0.3 | 506 |
| A | 2 | 8.1 | 574 | 3 weeks | 1.9 | 5 | <0.1 | 764 |
| A | 7 | 11.1 | 464 | 3 weeks | 1.8 | 6 | 0.1 | 470 |
| A | 9 | 5.6 | 439 | 3 weeks | 3.6 | 10 | <0.1 | 779 |
| A | 10 | 120.0 | 486 | 3 weeks | 28.6 | 3 | 1.0 | 657 |
| **A Avg (Range)** |  | **31.0 (5.6-120)** | **485 (439-574)** | **3 weeks** | **8.1 (1.8-28.6)** | **6.6 (3-10)** | **0.3 (<0.1-0.3)** | **635.2 (470-779)** |
| B | 3 | 7.2 | 512 | 6 weeks | 1.3 | 3 | 1.3 | 3 |
| B | 4 | 11.8 | 485 | 6 weeks | 2.8 | 3 | 0.8 | 3 |
| B | 5 | 8.3 | 389 | 6 weeks | 4 | 10 | 2.2 | 12 |
| B | 6 | 7.3 | 453 | 6 weeks | 9.8 | 3 | 2.4 | 12 |
| B | 8 | 39.4 | 160 | 6 weeks | 11.2 | 10 | 6.7 | 3 |
| **B Avg (Range)** |  | **14.9 (7.2-39.4)** | **400 (160-512)** | **6 weeks** | **5.8 (1.3-11.2)** | **5.8 (3-10)** | **2.7 (0.8-6.7)** | **6.6 (3-12)** |

For Cohort A, all patients underwent RARP/PLND prior to their third PSA measurement. Following RARP/PLND in Cohort A, PSA was undetectable in two patients and ≤0.3 ng/mL in three patients. All patients in Cohort A demonstrated a return to serum testosterone levels that were higher than their baseline values at three weeks after cessation of ADT. For Cohort B, one patient (Pt #6) showed an increase in PSA (7.3 ng/mL at baseline to 9.8 ng/mL after 3 weeks of ADT) before an ultimate decline of PSA to 2.4 ng/mL after 6 weeks of ADT. Testosterone remained in the castrate range (≤20 ng/dL) for Cohort B since they remained on ADT at the time of the third testosterone sample.

### Quantitative Evaluation of PSMA PET/CT (SUVmax)

The index lesion was evaluated as the region of interest (ROI) for all patients. The baseline SUVmax (prior to ADT) was compared to the post-ADT SUVmax for the same lesion. The percentage change in SUVmax was calculated (FIGURE 2).

**FIGURE 2.**
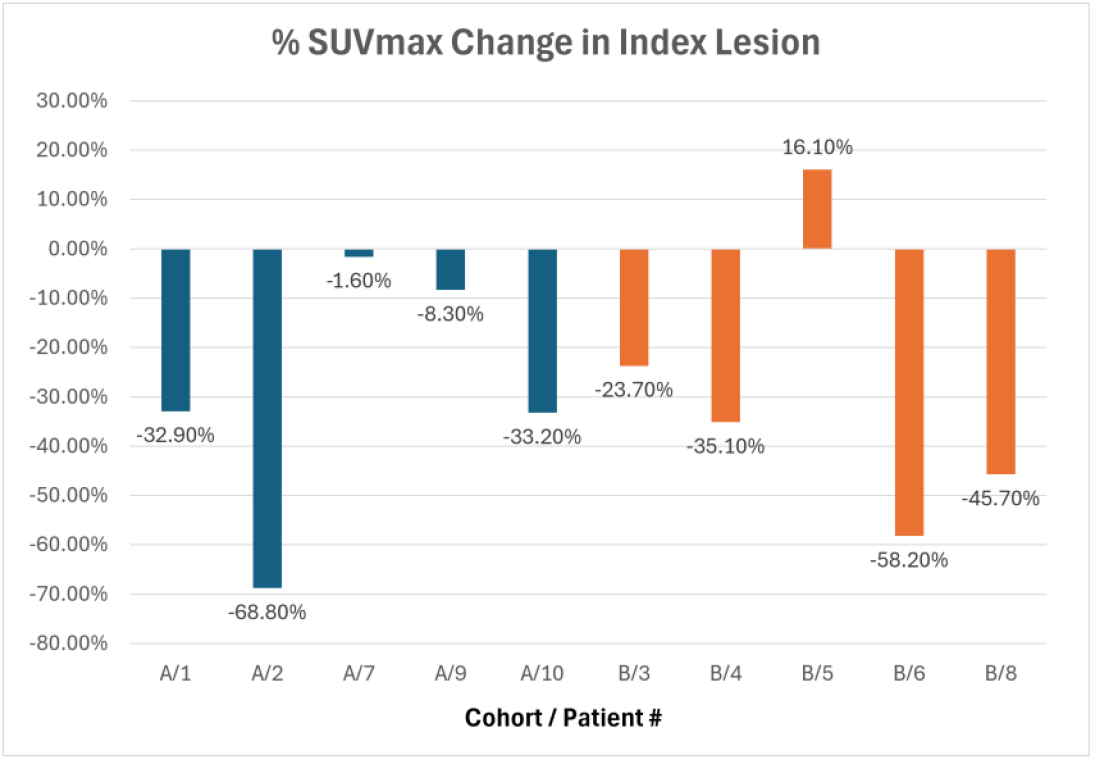
Cohort A (blue) 3 weeks of ADT, Cohort B (orange) 6 weeks of ADT. The %SUVmax change for the index lesion from pre-ADT PSMA PET/CT to post-ADT PSMA PET/CT for each patient

Rad1 and Rad2 concurred that one patient (Pt #5/Cohort B) had an increase in SUVmax of 16.1% in the index lesion after ADT (FIGURE 3).

**FIGURE 3.**
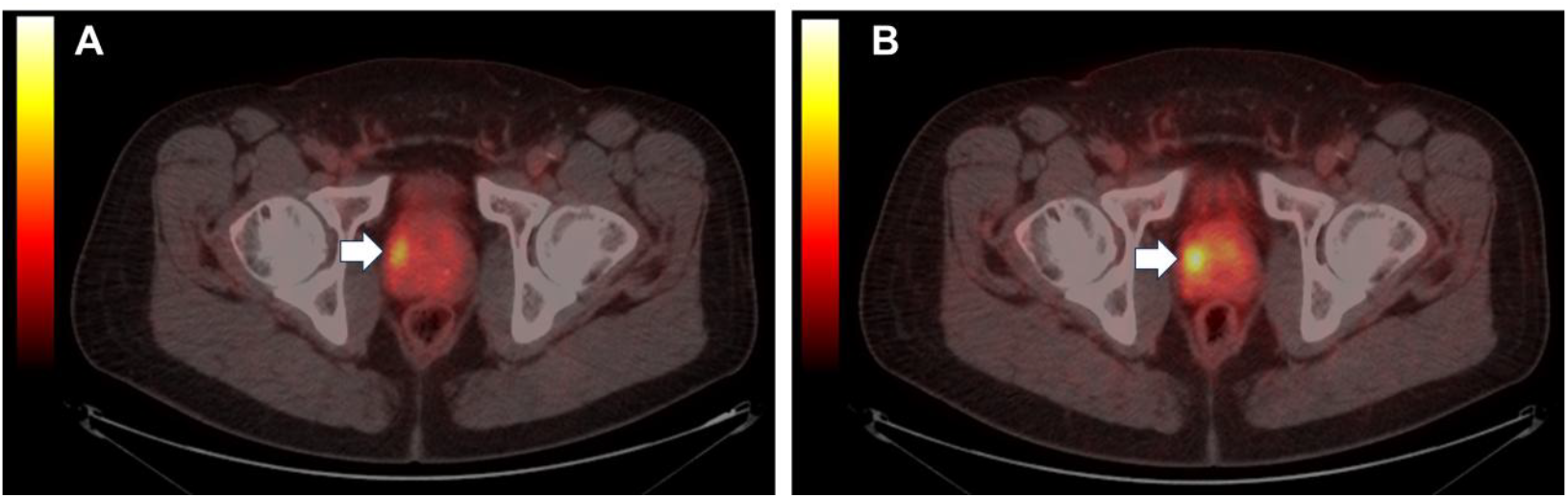
(A) Index lesion (arrow) in the right prostate prior to ADT. (B) Index lesion (arrow) in the right prostate with an increase in SUVmax after 6 weeks of ADT.

A decrease in SUVmax occurred in 9 of 10 patients (90%) after ADT (FIGURE 4). The agreement between Rad1 and Rad2 in determining SUVmax was 95% (differing by 0.1 unit of SUVmax on a single study) (Table 3).

**Table 3:** PSMA PET/CT results by evaluator demonstrating the change in SUVmax^*^ before and after ADT for the index lesion.

| Cohort | Pt ID | Blinded Nuclear Medicine Evaluation (Rad1) |  |  | Blinded Nuclear Medicine Evaluation (Rad2) |  |  |
| --- | --- | --- | --- | --- | --- | --- | --- |
|  |  | PSMA1 Index Lesion SUVmax | PSMA2 Index Lesion SUVmax | Diff. (% Diff.) | PSMA1 Index Lesion SUVmax | PSMA2 Index Lesion SUVmax | Diff. (% Diff.) |
| A | 1 | 7.9 | 5.3 | -2.6 (-32.9%) | 7.9 | 5.3 | -2.6 (-32.9%) |
| A | 2 | 16.0 | 5.0 | -11.0 (-68.8%) | 16.0 | 5.0 | -11.0 (-68.8%) |
| A | 7 | 6.1 | 6.0 | -0.1 (-1.6%) | 6.1 | 6.0 | -0.1 (-1.6%) |
| A | 9 | 3.6 | 3.3 | -0.3 (-8.3%) | 3.5 | 3.3 | -0.2 (-5.7%) |
| A | 10 | 36.1 | 24.1 | -12.0 (-33.2%) | 36.1 | 24.1 | -12.0 (-33.2%) |
| **A Avg** |  | **13.9** | **8.7** | **-5.2 (-29.0%)** | **13.9** | **8.7** | **-5.2 (-28.4%)** |
| B | 3 | 3.8 | 2.9 | -0.9 (-23.7%) | 3.8 | 2.9 | -0.9 (-23.7%) |
| B | 4 | 5.7 | 3.7 | -2.0 (-35.1%) | 5.7 | 3.7 | -2.0 (-35.1%) |
| B | 5 | 6.2 | 7.2 | 1.0 (16.1%) | 6.2 | 7.2 | 1.0 (16.1%) |
| B | 6 | 28.7 | 12.0 | -16.7 (-58.2%) | 28.7 | 12.0 | -16.7 (-58.2%) |
| B | 8 | 33.5 | 18.2 | -15.3 (-45.7%) | 33.5 | 18.2 | -15.3 (-45.7%) |
| **B Avg** |  | **15.6** | **8.8** | **-6.8 (-35.8%)** | **15.6** | **8.8** | **-6.8 (-35.8%)** |

\*SUVmax = $\frac{\text{maximum activity in the region of interest (mBq/g)}}{\text{(injected dose (mBq)/body weight (g))}}$

**FIGURE 4.**
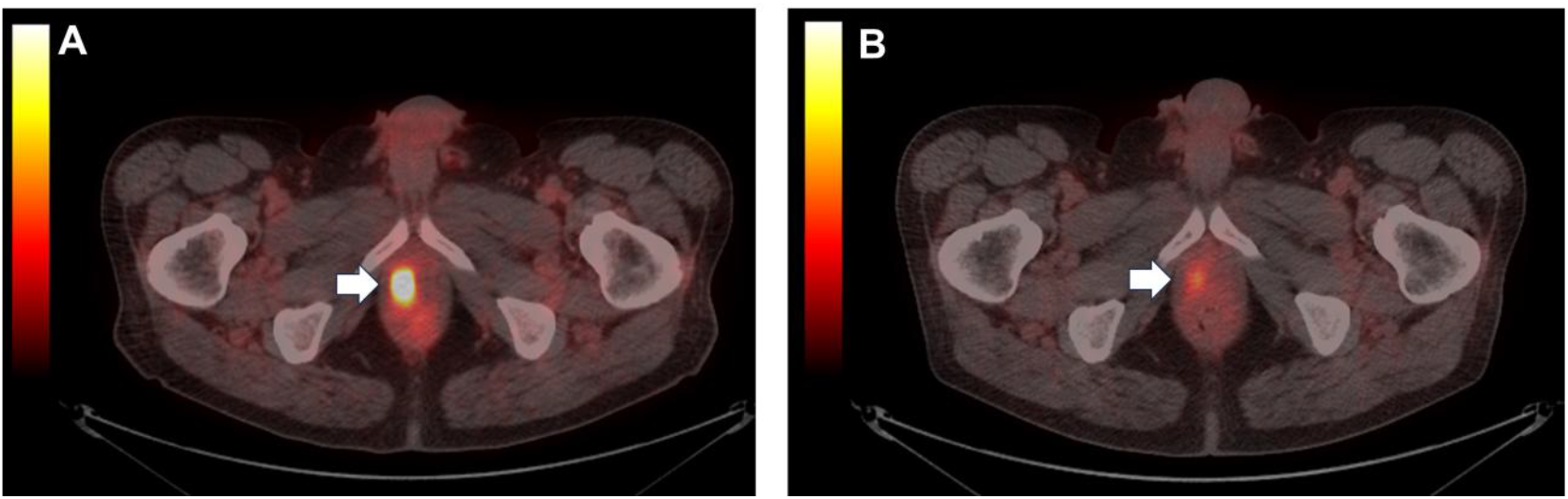
(A) Index lesion (arrow) prior to ADT. (B) Index lesion (arrow) with diminished SUVmax after 3 weeks of ADT.

### Qualitative Interpretation of PSMA PET/CT

The agreement between Rad1 and Rad2 for assigning an miTNM stage was 90%. There were minor disagreements within the staging system regarding the size and number of prostatic lesions, but none which would have changed clinical management. In one patient (Pt #6/Cohort B), there was some disagreement between radiologists about the number and location of positive lymph nodes (FIGURE 5). For another patient (Pt #10/Cohort A), one radiologist identified seminal vesicle involvement while the other did not.

**FIGURE 5.**
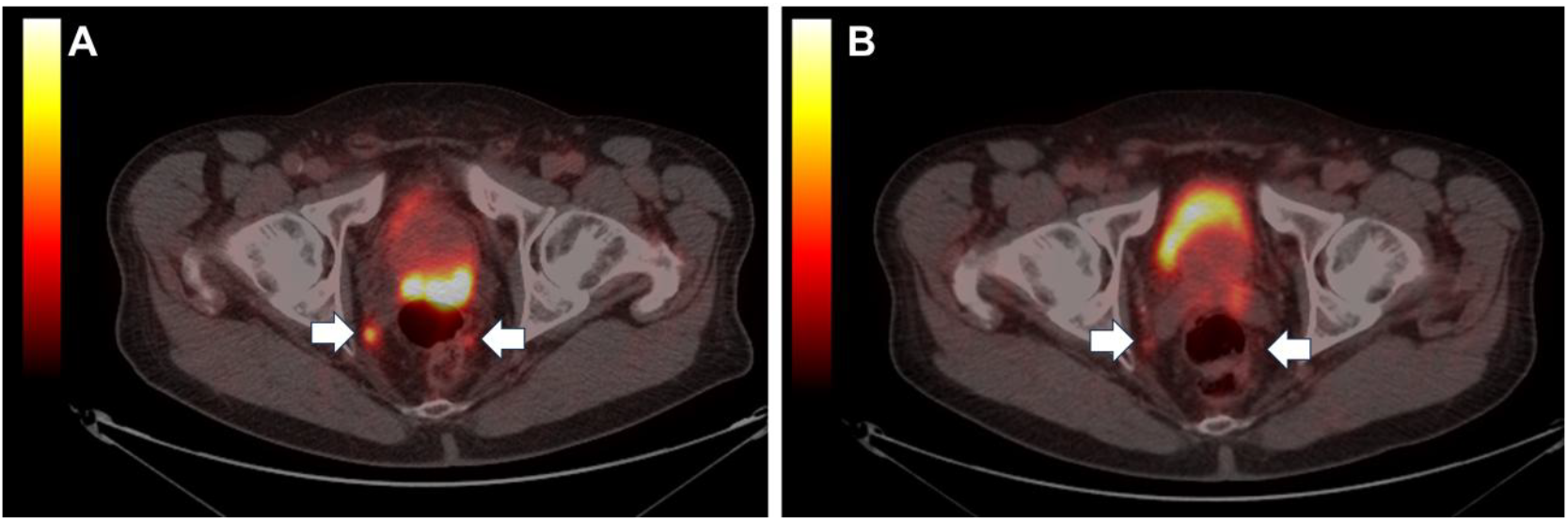
(A) Pre-ADT pelvic lymph nodes (arrows). (B) Post-ADT pelvic lymph nodes (arrows) less obvious.

There was some subjective change noted by the radiologists between PSMA PET/CT #1 and PSMA PET/CT #2 involving the visibility and number of intraprostatic lesions present. In two patients (Pt #6/Cohort B and Pt #10/Cohort A), with pathologic surgical stage T3b (seminal vesicle invasion) the stage was correctly predicted by PSMA PET/CT both before and after ADT. For 4 of 10 patients (40%), there was a change in miTNM stage from the pre-ADT to the post-ADT but none of the changes would have altered clinical management. There was no patient for which extraprostatic extension (EPE), seminal vesicle invasion, or nodal involvement seen on pre-ADT PSMA PET/CT was not seen on post-ADT PSMA PET/CT. Based on subsequent surgical pathology, PSMA PET/CT failed to demonstrate seminal vesicle invasion in one patient and incorrectly predicted seminal vesicle invasion in one patient.

### Surgical Pathology

All patients underwent RARP/PLND. No patient in either cohort had a positive surgical node. One patient in each Cohort had positive surgical margins and three patients in each Cohort had perineural invasion on final pathology. Three patients had EPE with negative surgical margins (one in Cohort A and two in Cohort B). One patient in each Cohort had seminal vesicle invasion (T3b). For the patient with an increase in SUVmax (Pt #5), final surgical pathology revealed a decrease in the Gleason score from 4+4=8 to 4+3=7. Four of five patients in each Cohort (80%) demonstrated a decrease in Gleason score from biopsy pathology to surgical pathology (Table 4).

**Table 4:**
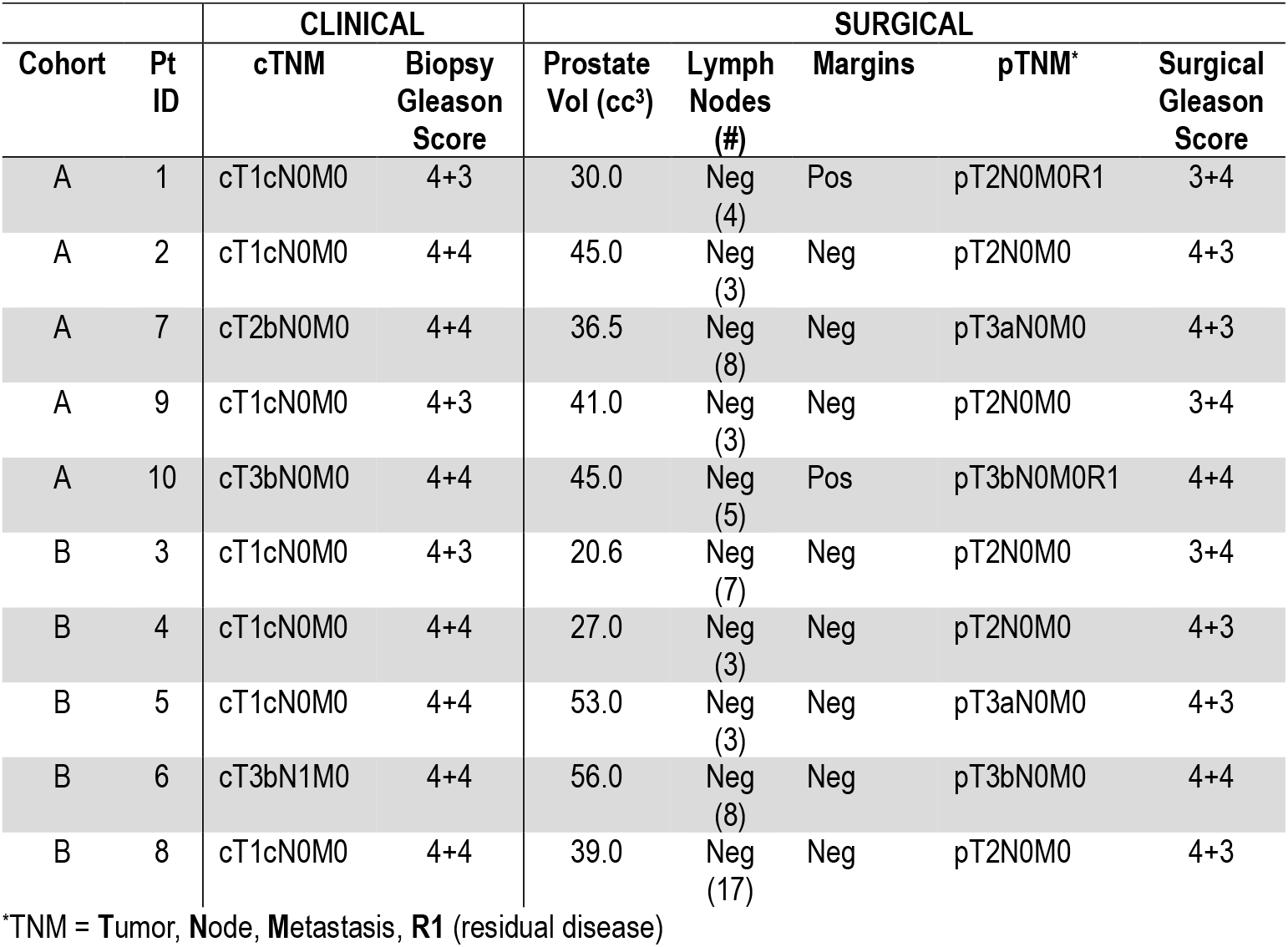
Clinical and surgical characteristics by Cohort.

## Discussion

There is increasing evidence that PSMA expression is androgen-repressed. Previous studies have shown that the initiation of ADT may result in the up-regulation of the FOLH1 gene, which encodes PSMA (thereby providing an explanation of the PSMA flare effect) in some patients with metastatic castrate-sensitive prostate cancer (mCSPC) and metastatic castrate-resistant prostate cancer (mCRPC) [4,5,8,9,10]. The manifestation of the PSMA flare effect was an increase in SUVmax on PSMA PET/CT. While the reported results were variable, there was an increase in SUVmax for 20-40% of patients which was most notable between 9 to 28 days of the initiation of ADT [5]. The heterogeneity of this response may be due to underlying features of the tumor itself. In the lesion-specific study of Malaspina, et al [9], 26% of metastatic bone metastases showed a 50% increase in SUVmax, 11% of lymph node metastases showed a 60% increase in SUVmax and 11% of primary prostate lesions showed a 45% increase in SUVmax at 3 to 4 weeks after the initiation of ADT. In the current study of patients undergoing PSMA PET/CT prior to RARP for localized prostate cancer, 1 of 10 (10%) patients demonstrated an increase in the SUVmax of the primary prostatic lesion on post-ADT PSMA PET/CT. All others demonstrated a decrease in the SUVmax of the primary prostatic lesion. The greater SUVmax decrease for Cohort B suggests that longer duration of ADT progressively suppresses PSMA expression.

It may be that primary, non-metastatic ADT-naïve lesions are less likely to manifest the flare effect than metastatic lesions (castrate-sensitive or castrate-resistant) owing to differences in the tumor microenvironment and androgen receptor signaling [11]. It is also possible that increased PSMA expression in index lesions had already peaked and begun to decline prior to both the 3-week (Cohort A) and 6-week (Cohort B) timing of the post-ADT PSMA PET/CT.

Though not the focus of this study, all patients treated with 3 weeks of ADT (Cohort A) had rapid and complete recovery of serum testosterone to pre-ADT levels by 21 days after discontinuation of ADT. There were no serious adverse events (SAE) reported for this study related to the use of relugolix. Hot flashes were the most frequently reported side-effect in 6 of 10 patients (60%), three in each cohort. Fatigue was reported by one patient who also reported hot flashes. Thus, the use of short-term ADT prior to RARP/PLND appears to be relatively safe with regards to the side effects of ADT.

There was a single patient suspected of having pelvic lymph node involvement (Pt #6/Cohort B) on PSMA PET/CT. Of note, the left obturator lymph node demonstrated a PSMA flare effect with an increase of SUVmax of 15.4%, (34.4 on the pre-ADT PSMA PET/CT and 39.7 on the post-ADT PSMA PET/CT). The index lesion for this patient did not show the flare effect. This left obturator lymph node was deemed by the surgeon to be unresectable based on involvement of the obturator nerve and therefore was not confirmed on surgical pathology. Additionally, the patient had two right external iliac nodes identified by both radiologists on the pre- and post-ADT PSMA PET/CT which were negative on surgical pathology. This likely represents a histopathologic treatment effect of ADT and highlights the importance of accounting for the use of neoadjuvant ADT when interpreting surgical pathology results.

There was no difference in the qualitative analysis of the post-ADT PSMA PET/CT scans which would have affected clinical management. Specifically, there was no clinical finding for any patient on pre-ADT imaging that was obscured or missed on post-ADT imaging. This suggests that short-term ADT (≤ 6 weeks) is unlikely to obscure important clinical findings of PSMA PET/CT. Our research is consistent with known variability in PSMA expression dynamics. Given the implications for future clinical trials and treatment pathways, further characterization and timing of ADT is critically important.

## Limitations

The small size and non-randomized nature of this pilot study precludes rigorous statistical analysis of the results. No control groups were included in this study. The timing of post ADT imaging may have missed an initial increased PSMA expression in some patients. The findings may not be generalizable to other PSMA PET/CT agents.

## Future Directions

Increased PSMA expression after ADT has the potential to increase the sensitivity of PSMA PET/CT for diagnostic and staging purposes but also for facilitating the identification of recurrent or residual disease at a lower serum PSA level for those patients with biochemical recurrence (BCR) [12]. Enhancing PSMA cellular expression at the time of PSMA-targeted radioligand therapy could possibly improve therapeutic effectiveness.

Direct or indirect quantitative, sequential measurement of PSMA in response to continuous ADT is needed to describe the timing, amplitude and duration of the PSMA flare effect. An indirect assessment of PSMA expression analysis by circulating tumor DNA, mRNA or gene-expression products could be considered. This would permit the non-invasive, frequent testing necessary to better characterize this effect, and better predict which patients are likely to experience and benefit from eliciting increased PSMA expression.

## Conclusion

Short-term ADT did not alter PSMA PET/CT findings in a manner that would impact surgical planning or management. No novel lesions (primary or metastatic) were detected on the PSMA PET/CT performed after ADT and no lesion present on pre-ADT imaging was obscured after ADT. The findings on this pilot study do not support the routine use of ADT prior to RARP/PLND to enhance PSMA PET/CT imaging; however, clinicians who use short-term ADT prior to RARP/PLND may do so with limited concern for lost information on a staging PSMA PET/CT.

## Data Availability

All data produced in the present study are available upon reasonable request to the authors

## Abbreviations

RARP: Robot-assisted radical prostatectomy
PLND: Pelvic lymphadenectomy
PSMA PET/CT: Prostate-specific membrane antigen positron emission tomography/computed tomography
PSMA: Prostate-specific membrane antigen
LHRH: Luteinizing hormone-releasing hormone
PSA: Prostate-specific antigen
EPE: Extraprostatic extension

## Disclosure

There are no conflicts of interest.

## Financial support

Blue Earth Diagnostics Ltd, Oxford, United Kingdom; Texas Health Resources Foundation, Arlington, TX; Texas Health Presbyterian Hospital Dallas, Dallas, TX

## Data Availability Statement

The data supporting the findings of this study are available from the corresponding author upon request.

## Author Contributions

PF Fulgham: protocol development, data analysis, manuscript writing

PW Barr: interpretation of PSMA PET/CT studies, manuscript editing

RR Bowman: interpretation PSMA PET/CT studies, manuscript editing

G Thoreson: manuscript editing

AR Clark: data collection, data analysis, manuscript editing

S Berger: protocol/project development, management of data quality

A Demitsas: management of patient recruitment and data collection

## Key Points

**Question:** Does the use of short-term ADT prior to PSMA PET/CT significantly enhance or diminish the findings with respect to clinical staging?

**Pertinent findings:** In two cohorts (n=5 in each) PSMA PET/CT followed by either 3 or 6 weeks of ADT and a second PSMA PET/CT were compared to determine if the change in SUVmax altered or enhanced clinically significant findings.

**Implications for patient care:** Short-term ADT (up to 6 weeks) does not significantly change the clinical findings on PSMA PET/CT used for initial staging of prostate cancer.

## Notes

### Competing Interest Statement

The authors have declared no competing interest.

### Clinical Trial

NCT07069465

### Funding Statement

This study was funded by Blue Earth Diagnostics Ltd, Oxford, United Kingdom; Texas Health Resources Foundation, Arlington, TX; Texas Health Presbyterian Hospital Dallas, Dallas, TX

### Author Declarations

Ethics committee/IRB of University of Texas Southwestern Medical Center gave ethical approval for this work

## References

1. Chow KM, So WZ, Lee HJ, et al. Head-to-head comparison of the diagnostic accuracy of prostate-specific membrane antigen positron emission tomography and conventional imaging modalities for initial staging of intermediate- to high-risk prostate cancer: a systematic review and meta-analysis. Eur Urol. 2023;84(1):36–48. 10.1016/j.eururo.2023.03.001

2. Hope TA, Eiber M, Armstrong WR, et al. Diagnostic accuracy of ^68^Ga-PSMA-11 PET for pelvic nodal metastasis detection prior to radical prostatectomy and pelvic lymph node dissection: a multicenter prospective phase 3 imaging trial. JAMA Oncol. 2021;7(11):1635–1642. 10.1001/jamaoncol.2021.3771

3. Ross AE, Yousefi K, Davicioni E, et al. Utility of risk models in decision making after radical prostatectomy: Lessons from a natural history cohort of intermediate- and high-risk men. Eur Urol. 2016; 69:496–504 10.1016/j.eururo.2015.04.016

4. Aggarwal R, Wei X, Kim W, et al. Heterogeneous flare in prostate specific membrane antigen positron emission tomography tracer uptake with initiation of androgen pathway blockade in metastatic prostate cancer. Eur Urol Oncol. 2018;1:78–82. 10.1016/j.euo.2018.03.010

5. Emmett L, Yin C, Crumbaker M, et al. Rapid modulation of PSMA expression by androgen deprivation: Serial ^68^Ga-PSMA-11 PET in men with hormone-sensitive and castrate-resistant prostate cancer commencing androgen blockade. J Nucl Med. 2019;60(7):950–950 10.2967/jnumed.118.223099

6. Heo, Y. Flotufolastat F 18: Diagnostic first approval. Mol Diagn Ther. 2023;27:631–636. 10.1007/s40291-023-00665-y

7. Eiber M, Herrmann K, Calais J, et al. Prostate cancer molecular imaging standardized evaluation (PROMISE): Proposed miTNM classification for the interpretation of PSMA-ligand PET/CT. J Nucl Med. 2018; 59:469–478 10.2967/jnumed.117.198119

8. Hope TA, Truillet C, Ehman EC, et al. ^68^Ga-PSMA-11 PET imaging of response to androgen receptor inhibition: first human experience. J Nucl Med. 2017;58:81–4. 10.2967/jnumed.116.181800

9. Malaspina S, Ettala O, Tolvanen T, et al. Flare on [^18^F]PSMA-1007 PET after short-term androgen deprivation therapy and its correlation to FDG uptake: possible marker of tumor aggressiveness in treatment-naïve metastatic prostate cancer patients. Eur J Nucl Med Mol Imaging. 2023;50:613–621. 10.1007/s00259-022-05970-y

10. Vaz S, Hadaschik B, Gabriel M, Herrmann K, Eiber M, Costa D. Influence of androgen deprivation therapy on PSMA expression and PSMA-ligand PET imaging of prostate cancer patients. Eur J Nucl Med Mol Imaging. 2020;47:9–15 (2020). 10.1007/s00259-019-04529-8

11. Bakht M, Beltran H. Biological determinants of PSMA expression, regulation and heterogeneity in prostate cancer. Nat Rev Urol. 2025;22:26–45. 10.1038/s41585-024-00900-z

12. Leitsmann C, Thelen P, Schmid M, et al. Enhancing PSMA-uptake with androgen deprivation therapy-a new way to detect prostate cancer metastases? Int Braz J Urol. 2019;45(3):459–467 10.1590/S1677-5538.IBJU.2018.0305

